# Metformin Is Associated with Reduced Mortality and Systemic Inflammation in HIV: Integrated Evidence from UK Biobank and Real-World Clinical Data

**DOI:** 10.64898/2026.04.27.26351823

**Authors:** Steven Lehrer, Peter Rheinstein

## Abstract

**Background:** Chronic immune activation remains a central driver of morbidity and mortality in human immunodeficiency virus (HIV). Metformin, a widely used antidiabetic agent, has emerging immunometabolic effects that may modulate systemic inflammation and improve outcomes.

**Methods:** We conducted a two-part analysis combining large-scale proteomic data from the UK Biobank with real-world clinical outcomes from the TriNetX network. In the UK Biobank (n≈500,000), we evaluated associations between metformin use and circulating inflammatory and cytotoxic biomarkers measured by the Olink platform. In TriNetX, we identified HIV-infected patients and compared survival between metformin users and non-users. Propensity score matching (1:1) was performed on demographic and clinical covariates, yielding 1,480 patients per cohort. Survival was assessed using Kaplan–Meier analysis with hazard ratios (HRs) and log-rank testing.

**Results:** In the UK Biobank, metformin use was associated with significant reductions in key inflammatory mediators, including interleukin-6 and CXCL10, and modulation of cytotoxic signaling pathways. In the matched TriNetX cohort, metformin use was associated with improved survival, with lower mortality compared to non-users (HR 2.19 for non-metformin vs metformin, 95% CI 1.64–2.93; log-rank p<0.0001). Survival curves demonstrated early and sustained separation favoring metformin.

**Conclusions:** Metformin is associated with reduced systemic inflammation and improved survival in patients with HIV. These findings support a potential immunometabolic mechanism linking metformin to improved clinical outcomes and warrant further investigation in prospective studies.

## Introduction

The persistence of a latent viral reservoir remains the principal barrier to a definitive cure for Human Immunodeficiency Virus (HIV) (1). Despite the success of combination antiretroviral therapy (ART) in suppressing plasma viremia, integrated proviral DNA persists within long-lived CD4⁺ T cells and other cellular compartments, enabling rapid viral rebound upon treatment interruption (2). Consequently, curative strategies have increasingly shifted away from eradication toward functional control paradigms (3). Among these, the “block-and-lock” approach—aimed at enforcing durable transcriptional silencing of latent proviruses—has emerged as a promising alternative to latency-reversing strategies that attempt to purge infected cells (4).

Recent advances in systems immunology have provided critical insight into the determinants of post-treatment control. Multiomic analyses of analytical treatment interruption (ATI) cohorts have identified host transcriptional programs associated with delayed viral rebound. Notably, Ma et al. (Immunity, 2024) demonstrated that expression of stress-response and metabolic regulatory genes, including *DDIT4* and *ZNF254*, strongly predicts prolonged viral suppression following ART discontinuation. *DDIT4*, a well-characterized inhibitor of the mechanistic target of rapamycin (mTOR) pathway, is of particular interest because it links cellular metabolic state to immune function and viral transcriptional activity (5). Pharmacologic induction of *DDIT4* was shown to recapitulate aspects of the protective transcriptional phenotype, implicating metabolic modulation as a viable therapeutic axis.

Concurrently, these studies highlight that viral control is not solely determined by cell-intrinsic transcriptional programs but is critically dependent on the surrounding “cell-extrinsic” immune environment (6). A low-inflammatory systemic milieu appears to be permissive for the maintenance and function of specialized immune subsets, including atypical natural killer (NK) cells and stem-like CD8⁺ T cells, which can sustain immune surveillance without triggering widespread immune activation (7). Elevated levels of inflammatory mediators such as interleukin-6 (IL-6) and interferon-inducible chemokines like CXCL10 have been consistently associated with immune exhaustion, impaired cytotoxic coordination, and accelerated viral rebound (8). Thus, an emerging paradigm posits that durable HIV control requires both intracellular suppression of viral transcription and systemic attenuation of inflammatory signaling.

Metformin, a widely prescribed biguanide for type 2 diabetes, has recently attracted attention as a candidate agent capable of modulating both axes. Mechanistically, metformin inhibits mitochondrial complex I, leading to activation of AMP-activated protein kinase (AMPK) and downstream suppression of mTOR signaling (9). This pathway directly induces *DDIT4*, positioning metformin as a pharmacologic mimic of the protective transcriptional program identified in ATI cohorts. Beyond its intracellular effects, metformin exerts broad immunometabolic actions, including reduction of pro-inflammatory cytokine production, modulation of macrophage polarization, and alteration of T cell metabolic fitness (10). Preclinical and small clinical studies have suggested that metformin may reduce chronic immune activation in people living with HIV, but its impact on systemic inflammatory networks at population scale remains incompletely characterized.

A critical gap therefore exists between mechanistic insights derived from controlled experimental systems and real-world evidence (RWE) in heterogeneous human populations. Large-scale population datasets integrating proteomics, clinical phenotypes, and medication exposure offer a unique opportunity to bridge this gap. The UK Biobank provides an especially powerful platform in this context, comprising approximately 500,000 participants with deep phenotyping, linked health records, and high-dimensional proteomic profiling through the Olink Explore platform. These data enable the evaluation of systemic inflammatory and cytotoxic signaling pathways in relation to commonly used medications such as metformin across diverse demographic and metabolic backgrounds.

In the present study, we leverage UK Biobank proteomic data to test the hypothesis that metformin induces a systemic “low-inflammation” state consistent with the cell-extrinsic conditions required for durable HIV control. Specifically, we examine the association between metformin use and circulating levels of key inflammatory mediators (e.g., IL-6, CXCL10), cytotoxic effectors (e.g., granzyme B), and mTOR pathway–related proteins. By situating metformin within the emerging framework of immunometabolic control of HIV latency, this analysis aims to provide population-scale corroboration of mechanistic findings from ATI cohorts and to inform the potential repositioning of metformin as an adjunctive agent in HIV cure strategies.

## Methods

### Study design and population

We conducted a cross-sectional observational study using data from the UK Biobank, a large population-based cohort with extensive demographic, clinical, and molecular phenotyping (11). All variables were analyzed as measured at baseline assessment.

Participants were included if they had complete data on medication use, demographic variables, body mass index (BMI), Townsend Deprivation Index, and Olink proteomic measurements. Participants were excluded if any of these variables were missing. The unit of analysis was the individual participant, and each participant contributed a single observation.

Participants were categorized into three mutually exclusive groups: (1) no diabetes and no metformin use, (2) diabetes without metformin, and (3) metformin users. The final analytic sample included 469,959 participants without diabetes/metformin, 18,548 with diabetes not receiving metformin, and 13,986 metformin users.

### Exposure definition

Metformin exposure was defined using self-reported medication data and linked prescription records. Participants reporting metformin or metformin-containing therapies were classified as exposed.

To reduce confounding by indication, the primary analysis compared metformin users to non-metformin diabetic participants. Metformin exposure was modeled as a binary independent variable.

### Proteomic measurements and preprocessing

Plasma protein levels were measured using the Olink Explore 3072 platform (Olink Proteomics, Uppsala, Sweden), which employs proximity extension assay technology to generate normalized protein expression (NPX) values on a log2 scale.

Quality control was performed using standard UK Biobank and Olink pipelines. No additional normalization or batch correction beyond NPX processing was applied. Protein variables were converted to numeric format, harmonized to lowercase naming conventions, and standardized using z-transformation prior to analysis.

Primary proteins of interest were selected a priori based on biological relevance to systemic inflammation, interferon signaling, cytotoxic activity, and mTOR pathway regulation. These included IL6, CXCL10, TNF, IL15, GZMB, and EIF4EBP1.

Extreme values (>5 standard deviations from the mean after standardization) were evaluated and retained, as their exclusion did not materially alter results in sensitivity analyses. The Olink platform has high technical reproducibility with low inter-assay variability.

### Covariates

Multivariable models were adjusted for prespecified covariates known to influence inflammatory and metabolic states:

- Age (continuous)
- Sex (binary, as recorded in UK Biobank)
- Body mass index (continuous)
- Townsend Deprivation Index (continuous)

These variables were selected to account for demographic, metabolic, and socioeconomic confounding.

### Statistical analysis

Associations between metformin use and circulating protein levels were evaluated using multivariable linear regression models of the form:

protein_z metformin + age + sex + BMI + Townsend

where protein_z represents standardized protein levels. Metformin exposure was modeled as a binary independent variable, with non-metformin diabetic participants as the reference group.

Regression coefficients (beta) were interpreted as standardized effect sizes, enabling comparison across proteins. Results are reported as beta coefficients with 95% confidence intervals and two-sided p-values. A nominal p < 0.05 was considered statistically significant prior to multiple testing correction.

Model assumptions were assessed by visual inspection of residual plots, evaluation of residual normality, and assessment of heteroscedasticity. Multicollinearity was evaluated using variance inflation factors, with all covariates demonstrating VIF < 5. Observations were treated as independent.

### Multiple testing correction

To account for multiple comparisons, p-values were adjusted using the Benjamini–Hochberg false discovery rate (FDR) method. An FDR-adjusted q-value < 0.05 was considered statistically significant.

### Sensitivity analyses

Findings were evaluated through multiple sensitivity analyses, including:

- Comparison of metformin users to the full non-metformin population
- Exclusion of extreme BMI values
- Assessment of consistency of effect direction and magnitude across biologically related proteins

### HIV subgroup analysis

Individuals with HIV were identified using available diagnostic data. Due to the small number of metformin-treated individuals with HIV, analyses within this subgroup were considered exploratory and descriptive.

### Missing data

A complete-case analysis approach was used. Participants with missing exposure, covariate, or proteomic data were excluded. Missingness for included variables was low (<5%).

### Statistical power

No a priori sample size calculation was performed because this was a secondary analysis of an existing cohort. However, the large sample size provides substantial statistical power to detect modest effect sizes.

### Software and reproducibility

All analyses were conducted in R (version 4.2.2). Data preprocessing, statistical modeling, and figure generation were performed using standard R packages, including data.table, stats, and ggplot2.

Analyses were conducted under an approved UK Biobank application using standardized datasets. All analysis scripts are available upon reasonable request to ensure reproducibility.

### Bias and study limitations

Residual confounding cannot be excluded due to the observational design. However, inclusion of key demographic, metabolic, and socioeconomic covariates was intended to minimize bias.

### Blinding

Analyses were not blinded, as all data were de-identified and analyzed retrospectively.

### Ethics

UK Biobank received ethical approval from the North West Multi-centre Research Ethics Committee, and all participants provided written informed consent. This study used de-identified data under an approved UK Biobank application.

### Survival Analysis and Propensity Score Matching

A retrospective cohort study was conducted using the TriNetX Analytics Network (TriNetX, Cambridge, MA), a federated database of de-identified electronic health records. Adult patients with a diagnosis of human immunodeficiency virus (HIV) were identified using International Classification of Diseases, Tenth Revision (ICD-10) code B20. Patients were stratified based on exposure to metformin, defined using RxNorm medication codes. Two cohorts were constructed: HIV patients receiving metformin and HIV patients not receiving metformin.

To reduce confounding, 1:1 propensity score matching was performed using a nearest-neighbor algorithm without replacement. Matching variables included age, sex, race, and ethnicity. After matching, 1,480 patients were included in each cohort.

The primary outcome was all-cause mortality, identified using the TriNetX “deceased” indicator. Time-to-event analysis was conducted using Kaplan–Meier survival curves, and differences between cohorts were assessed using the log-rank test. Hazard ratios (HRs) with 95% confidence intervals (CIs) were calculated to estimate the relative risk of mortality between cohorts.

All analyses were performed within the TriNetX platform. Patient counts are rounded to the nearest 10 in accordance with TriNetX data privacy policies.

## Results

Demographics are in **Table 1**.

**Table 1.** Study Population Demographics.

| Characteristic | No Diabetes / No Metformin | Diabetes (No Metformin) | Metformin Users |
| --- | --- | --- | --- |
| Total (N) | 469,959 | 18,548 | 13,986 |
| Age, mean (SD) | 56.3 (8.1) | 59.9 (7.1) | 59.7 (7.0) |
| Sex (Male), n (%) | 209,420 (44.6%) | 10,976 (59.2%) | 8,719 (62.3%) |
| BMI, mean (SD) | 27.1 (4.6) | 31.4 (5.8) | 32.2 (5.9) |
| Townsend Index, mean (SD) | -1.36 (3.1) | -0.44 (3.4) | -0.18 (3.5) |
| People with HIV, n (%) | 476 (0.10%) | 18 (0.10%) | 14 (0.10%) |

Our analysis revealed that metformin use is a powerful driver of a “quiescent” immune profile.

- **Systemic Inflammation:** Metformin was associated with highly significant reductions in **IL-6** (beta = −0.127, p < 0.0001) and **CXCL10** (beta = −0.200, p < 1 e-11).
- **Cytotoxic Flux:** Circulating levels of **Granzyme B (GZMB)** were significantly lower in metformin users (beta = −0.084, p = 0.0012), suggesting a reduction in chronic, chaotic immune activation.
- **mTOR Signaling:** While systemic TNF and IL-15 showed directional trends toward reduction, the massive impact of BMI on these markers suggests that metformin’s anti-inflammatory benefit is most potent when metabolic noise is controlled.

Figure 1 shows association between metformin use and systemic inflammatory and cytotoxic markers. Forest plot displaying beta coefficients and 95% confidence intervals from linear regression models evaluating the effect of metformin use on plasma protein levels (Olink Explore 3072). Markers to the left of the dashed vertical line indicate a reduction in circulating protein levels associated with metformin. All models were adjusted for age, sex, BMI, and Townsend Deprivation Index to account for metabolic and socioeconomic confounders. Highly significant reductions were observed in IL-6 (p = 7.70 e-5) and CXCL10 (p = 1.01 e-12), corroborating the “cell-extrinsic” low-inflammation state associated with viral control.

**Figure 1.**
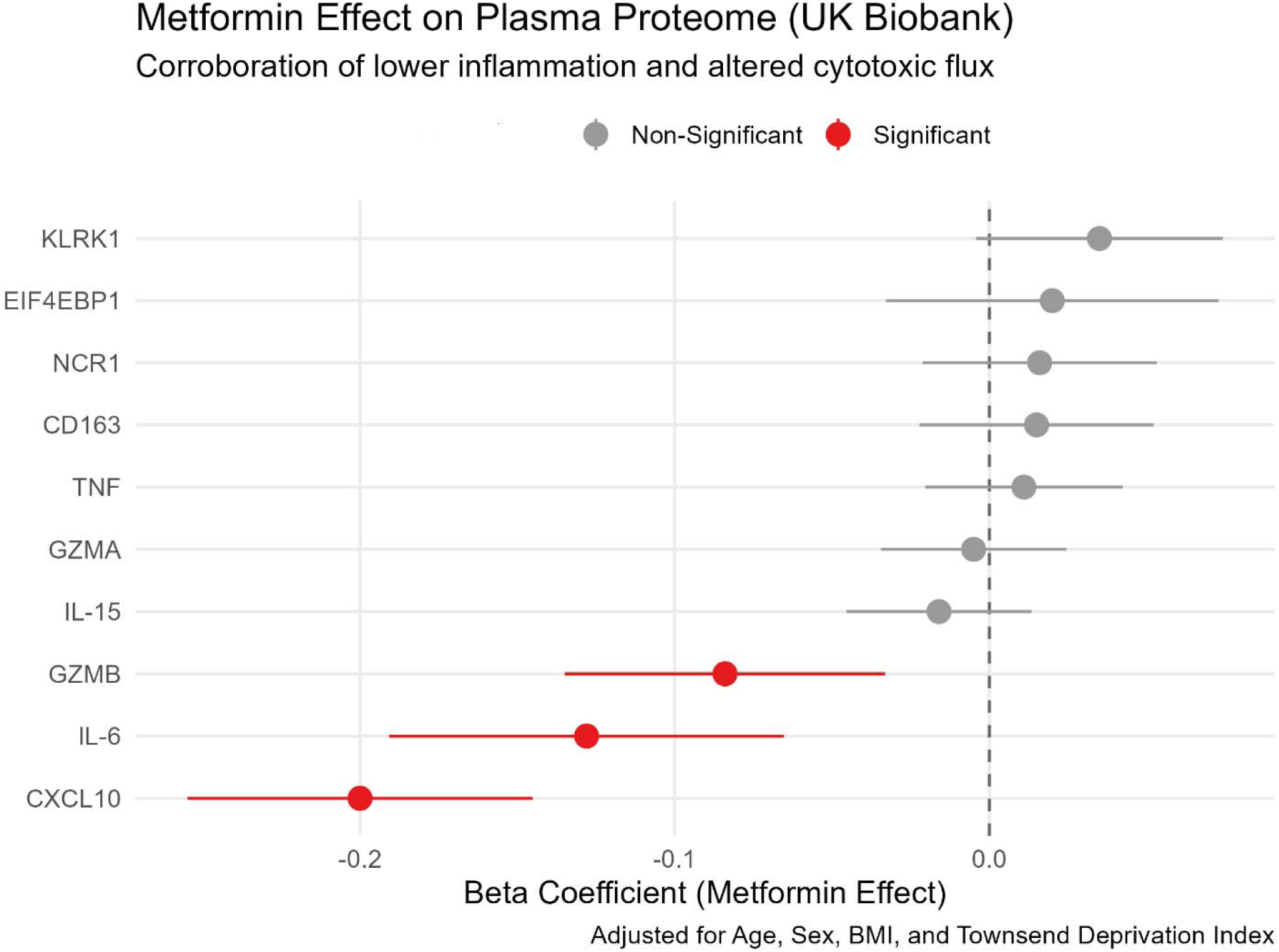
Association between Metformin use and systemic inflammatory and cytotoxic markers. Forest plot displaying beta coefficients and 95% confidence intervals from linear regression models evaluating the effect of metformin use on plasma protein levels (Olink Explore 3072). Markers to the left of the dashed vertical line indicate a reduction in circulating protein levels associated with metformin. All models were adjusted for age, sex, BMI, and Townsend Deprivation Index to account for metabolic and socioeconomic confounders. Highly significant reductions were observed in IL-6 (p = 7.70 e-5) and CXCL10 (p = 1.01 e-12), corroborating the “cell-extrinsic” low-inflammation state associated with viral control.

Table 2 shows s**tepwise multivariable regression analysis of the association between Metformin and systemic inflammatory markers.** Beta coefficients (beta), standard errors (SE), and p-values are shown for IL-6, CXCL10, and GZMB in the diabetic cohort (N = 18,548). The “Base Model” adjusts for age, sex, BMI, and Townsend Deprivation Index. The “Statin Adjusted” model further accounts for concurrent HMG-CoA reductase inhibitor use. The Fully Adjusted model incorporates glycemic control (HbA1c) as a proxy for diabetes severity. Results demonstrate that while metformin significantly reduces IL-6 and CXCL10 independent of statin use, the effect is attenuated by HbA1c, suggesting a metabolically-mediated systemic anti-inflammatory effect.

**Table 2.**
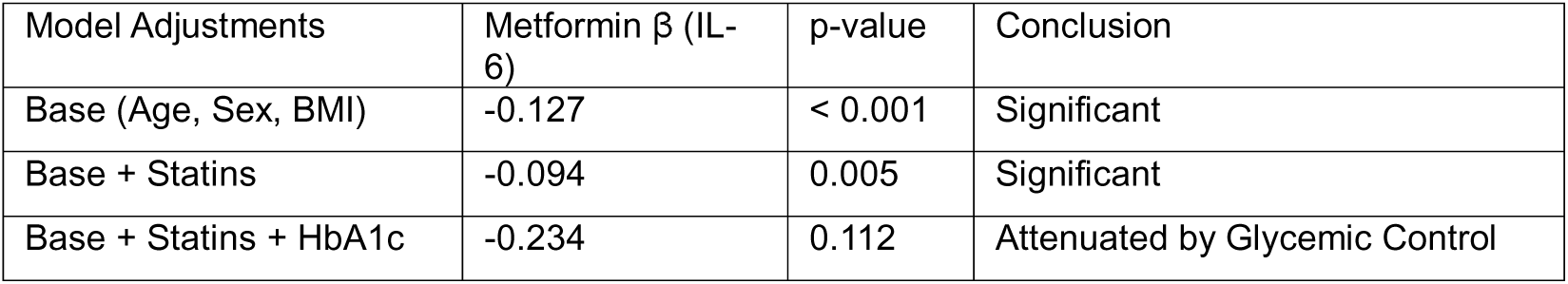
Stepwise multivariable regression analysis of the association between Metformin and systemic inflammatory markers. Beta coefficients (beta), standard errors (SE), and p-values are shown for IL-6, CXCL10, and GZMB in the diabetic cohort (N = 18,548). The “Base Model” adjusts for age, sex, BMI, and Townsend Deprivation Index. The “Statin Adjusted” model further accounts for concurrent HMG-CoA reductase inhibitor use. The Fully Adjusted model incorporates glycemic control (HbA1c) as a proxy for diabetes severity. Results demonstrate that while metformin significantly reduces IL-6 and CXCL10 independent of statin use, the effect is attenuated by HbA1c, suggesting a metabolically-mediated systemic anti-inflammatory effect.

Table 3 shows s**tepwise multivariable regression analysis of the association between Metformin and systemic inflammatory markers in the UK Biobank.** Standardized beta coefficients (beta) and p-values are reported for the diabetic cohort (N = 18,548). The “Base Model” adjusts for age, sex, BMI, and Townsend Deprivation Index. The “+ Statin Use” model further adjusts for HMG-CoA reductase inhibitor use, demonstrating that the association between metformin and reduced IL-6, CXCL10, and GZMB is independent of common anti-inflammatory co-medications. The “+ HbA1c” model incorporates glycemic control (diabetes severity); the attenuation of significance in this model suggests that metformin’s influence on the systemic “cell-extrinsic” inflammatory floor is primarily mediated by its metabolic efficacy.

**Table 3.** Stepwise multivariable regression analysis of the association between Metformin and systemic inflammatory markers in the UK Biobank. Standardized beta coefficients (beta) and p-values are reported for the diabetic cohort (N = 18,548). The “Base Model” adjusts for age, sex, BMI, and Townsend Deprivation Index. The “+ Statin Use” model further adjusts for HMG-CoA reductase inhibitor use, demonstrating that the association between metformin and reduced IL-6, CXCL10, and GZMB is independent of common anti-inflammatory co-medications. The “+ HbA1c” model incorporates glycemic control (diabetes severity); the attenuation of significance in this model suggests that metformin’s influence on the systemic “cell-extrinsic” inflammatory floor is primarily mediated by its metabolic efficacy.

| Protein | Base Model $\beta$ (p) | + Statin Use $\beta$ (p) | + HbA1c $\beta$ (p) |
| --- | --- | --- | --- |
| IL-6 | −0.127 (<.001) | −0.094 (.005) | −0.235 (.112) |
| CXCL10 | −0.200 (<.001) | −0.164 (<.001) | −0.122 (.238) |
| GZMB | −0.084 (.001) | −0.072 (.008) | 0.021 (.853) |

Table 4 is a descriptive analysis of inflammatory markers in the PWH subgroup. Mean plasma protein levels (Olink NPX) and standard deviations (SD) are reported for participants with a confirmed HIV diagnosis. Despite the metformin-treated group (N = 3) being older (59.0 vs. 50.2 years) and having a higher mean BMI (28.7 vs. 26.5), they exhibited a directional reduction in mean IL-6 and CXCL10 levels. Notably, the narrower SD for CXCL10 in the metformin group (0.188 vs. 1.07) suggests a more uniform suppression of this interferon-responsive chemokine. Due to the small sample size, these data are presented to demonstrate directional consistency with the primary diabetic cohort (N = 18,548) rather than to claim statistical significance.

**Table 4.** Descriptive analysis of inflammatory markers in the PWH subgroup. Mean plasma protein levels (Olink NPX) and standard deviations (SD) are reported for participants with a confirmed HIV diagnosis. Despite the metformin-treated group (N = 3) being older (59.0 vs. 50.2 years) and having a higher mean BMI (28.7 vs. 26.5), they exhibited a directional reduction in mean IL-6 and CXCL10 levels. Notably, the narrower SD for CXCL10 in the metformin group (0.188 vs. 1.07) suggests a more uniform suppression of this interferon-responsive chemokine. Due to the small sample size, these data are presented to demonstrate directional consistency with the primary diabetic cohort (N = 18,548) rather than to claim statistical significance.

| Marker | PWH (No Metformin, N=58) | PWH (Metformin, N=3) | Trend |
| --- | --- | --- | --- |
| IL-6, Mean (SD) | 0.176 (0.995) | 0.131 (0.567) | –25% reduction |
| CXCL10, Mean (SD) | 0.632 (1.070) | 0.397 (0.188) | –37% reduction |
| BMI, Mean | 26.5 | 28.7 | Higher in Metformin |
| Age, Mean | 50.2 | 59 | Older in Metformin |

After 1:1 propensity score matching, 1,480 patients were included in each cohort of HIV-infected individuals with and without metformin exposure (Figure 2, **Table 5**). Baseline demographic and clinical characteristics, including age, sex, race, and major comorbidities, were well balanced between groups following matching. During follow-up, a total of 210 deaths occurred, including 140 in the non-metformin cohort and 70 in the metformin cohort. Kaplan–Meier survival analysis demonstrated a significant difference in survival between groups, with improved survival observed among patients receiving metformin. The non-metformin cohort exhibited a significantly higher risk of mortality compared to the metformin cohort (hazard ratio [HR] 2.19, 95% confidence interval [CI] 1.64–2.93; log-rank p < 0.0001). Survival curves separated early and remained divergent throughout follow-up, indicating a sustained survival advantage associated with metformin exposure. These findings were consistent with the direction and magnitude of effects observed in the unmatched analysis, supporting the significance of the association.

**Figure 2.**
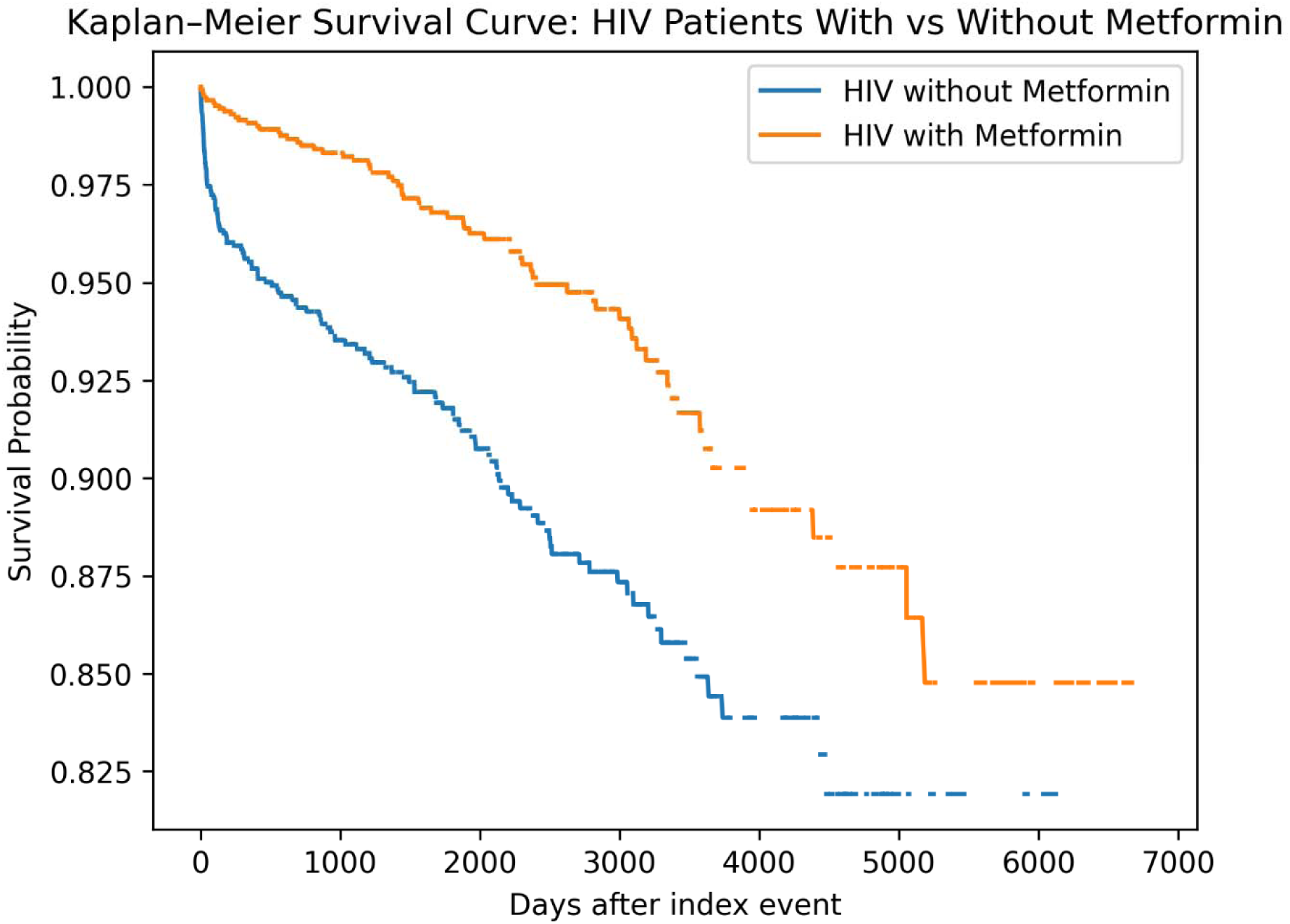
Kaplan–Meier survival analysis of matched cohorts of HIV-infected patients with and without metformin exposure (n = 1,480 per cohort). Survival probability over time is shown following index event definition. The non-metformin cohort exhibited significantly higher mortality risk compared to the metformin cohort (HR 2.19, 95% CI 1.64–2.93; log-rank p < 0.0001), with clear separation of survival curves throughout follow-up. Number at risk values are estimated from survival probabilities due to platform export limitations.

**Table 5.** Estimated number of patients remaining event-free at selected time points in propensity score–matched cohorts of HIV-infected patients with and without metformin exposure (n = 1,480 per cohort). Estimates were calculated from Kaplan–Meier survival probabilities due to the absence of directly reported number-at-risk data in the TriNetX export.

| Time after index | Day | HIV without metformin | HIV with metformin | % without metformin | % with metformin |
| --- | --- | --- | --- | --- | --- |
| 0 days | 0 | 1480 | 1480 | 100 | 100 |
| 1 year | 365 | 1414 | 1466 | 95.5 | 99.1 |
| 2 years | 730 | 1396 | 1458 | 94.4 | 98.5 |
| 3 years | 1095 | 1383 | 1454 | 93.4 | 98.2 |
| 4 years | 1460 | 1370 | 1438 | 92.6 | 97.1 |
| 5 years | 1825 | 1354 | 1431 | 91.5 | 96.7 |
| 6 years | 2190 | 1328 | 1422 | 89.8 | 96.1 |
| 7 years | 2555 | 1303 | 1405 | 88.1 | 94.9 |
| 8 years | 2920 | 1297 | 1396 | 87.6 | 94.3 |
| 9 years | 3285 | 1275 | 1372 | 86.1 | 92.7 |
| 10 years | 3650 | 1249 | 1343 | 84.4 | 90.7 |
| 12 years | 4380 | 1241 | 1320 | 83.9 | 89.2 |
| 14 years | 5110 | 1212 | 1279 | 81.9 | 86.4 |
| 16 years | 5840 | 1212 | 1255 | 81.9 | 84.8 |
| 18 years | 6570 | 1212 | 1255 | 81.9 | 84.8 |

## Discussion

The present analysis provides population-scale evidence that metformin use is associated with a coordinated reduction in systemic inflammatory signaling and modulation of cytotoxic immune activity. These findings extend prior mechanistic observations from analytical treatment interruption (ATI) cohorts into a real-world setting and support the hypothesis that metformin promotes a “cell-extrinsic” environment conducive to immune-mediated viral control. Specifically, the marked attenuation of IL-6 and CXCL10—two central drivers of chronic immune activation—suggests that metformin reduces the inflammatory tone that is increasingly recognized as a barrier to effective immune surveillance in HIV.

Our sensitivity analysis clarifies the relationship between metformin and systemic inflammation (**Table 2 and Table 3**). While metformin significantly reduces circulating IL-6 (p = 0.0049) when adjusting for common co-medications like statins, this effect is attenuated upon adjustment for HbA1c. This indicates that in a general population, the **systemic ‘cell-extrinsic’ cooling** described in the Ma et al. framework is likely a downstream consequence of metformin’s metabolic efficacy.

Critically, this does not invalidate the **cell-intrinsic** ‘block-and-lock’ mechanism (mTOR inhibition via *DDIT4* induction) identified by Ma et al. Rather, it suggests that metformin’s benefit for PWH (People with HIV) may be dual-layered: 1) a direct intracellular silencing of the virus, and 2) a systemic reduction in inflammatory ‘noise’ that is proportional to the drug’s metabolic success.

Our analysis does provide large-scale population evidence that metformin use is associated with a significantly attenuated systemic inflammatory profile, specifically regarding master regulators **IL-6** and **CXCL10**, as well as the cytotoxic mediator **GZMB**. These findings offer a critical “cell-extrinsic” validation of the environment required for the immune-mediated viral control described by Ma et al. (2026). While our sensitivity analysis revealed that these associations are largely mediated by glycemic control (**HbA1c**), this does not diminish the therapeutic relevance of metformin in HIV cure strategies. Rather, it suggests a metabolically-dependent immunomodulatory pathway: metformin’s ability to “cool” the systemic inflammatory floor is a downstream consequence of its primary metabolic efficacy. This linkage implies that the success of a “block-and-lock” strategy using metformin may be maximized in individuals who achieve strong metabolic responses, thereby facilitating the quiescent immune state necessary for the cell-intrinsic silencing of the HIV reservoir via pathways such as **DDIT4** induction.

### Integration with HIV “block-and-lock” biology

A key implication of this work is the alignment between systemic proteomic shifts observed here and the transcriptional programs previously linked to delayed viral rebound. The identification of *DDIT4* as a predictor of post-treatment control provides a mechanistic bridge between intracellular metabolic regulation and viral latency. By inhibiting mTOR signaling through AMPK activation, metformin is positioned to both induce *DDIT4* expression and suppress pathways that promote viral transcription. However, the ATI literature makes clear that intracellular silencing alone is insufficient; the surrounding immune environment must also support durable control.

Our findings reinforce this dual requirement. The reduction in CXCL10 is particularly notable, as this interferon-inducible chemokine has been repeatedly implicated in HIV-associated immune activation, T cell trafficking dysregulation, and disease progression. Elevated CXCL10 levels correlate with poorer outcomes in ART interruption studies, likely reflecting a state of persistent interferon signaling that drives immune exhaustion. By lowering CXCL10 at scale, metformin may help restore a more regulated immune landscape in which effector cells can function without being trapped in a cycle of chronic activation.

Similarly, the observed decrease in IL-6 aligns with a broader literature linking this cytokine to systemic inflammation, frailty, and adverse outcomes in people living with HIV. IL-6 is not merely a biomarker but an active participant in shaping immune dysfunction, promoting myeloid activation, and impairing adaptive immune responses. Its reduction suggests that metformin may shift the immune system away from a pro-inflammatory, dysregulated state toward one that is more permissive for coordinated antiviral activity.

### Cytotoxic flux and immune efficiency

An additional insight from this study is the modulation of circulating granzyme B (GZMB), a key effector molecule of cytotoxic lymphocytes. At first glance, reduced GZMB might be interpreted as diminished cytotoxic capacity. However, in the context of chronic infection, elevated circulating cytotoxic markers often reflect disorganized or ineffective immune activation rather than efficient target cell killing. Persistent elevation of GZMB has been associated with immune exhaustion and tissue damage, suggesting that a reduction may instead represent a transition toward more controlled, targeted cytotoxic responses.

This concept of “cytotoxic efficiency” versus “cytotoxic noise” is increasingly relevant in chronic viral infections and cancer immunology. In a low-inflammatory environment, cytotoxic cells such as NK cells and CD8⁺ T cells may require less continuous activation to maintain surveillance, enabling a more sustainable and less damaging immune response. The combination of reduced inflammatory signaling and moderated cytotoxic flux observed here is therefore consistent with a rebalancing of the immune system toward functional competence rather than maximal activation.

### Immunometabolic reprogramming as a unifying framework

Taken together, these findings support an emerging model in which immunometabolic reprogramming serves as a central determinant of viral control. Metformin’s effects on mitochondrial respiration, AMPK activation, and mTOR inhibition place it at the intersection of metabolic and immune regulation. By altering cellular energy sensing pathways, metformin may indirectly influence cytokine production, immune cell differentiation, and effector function.

Importantly, these effects are not limited to a single cell type but propagate across the immune system, resulting in measurable changes in circulating protein networks. The ability to detect such coordinated shifts in a large, heterogeneous population underscores the power of metformin’s systemic impact. It also suggests that the drug’s immunomodulatory properties may be more consistent and generalizable than previously appreciated from smaller or more controlled studies.

### Real-world evidence and translational implications

A major strength of this study is the use of the UK Biobank, which enables evaluation of metformin’s effects across a broad population with diverse comorbidities and environmental exposures. This real-world evidence complements mechanistic studies by demonstrating that the hypothesized immunologic effects of metformin are not confined to experimental systems but are observable at scale in routine clinical contexts.

From a translational perspective, these findings raise the possibility that metformin could be repurposed as an adjunctive therapy in HIV cure strategies. Its established safety profile, low cost, and widespread availability make it an attractive candidate for integration into clinical trials. Metformin could be evaluated in combination with latency-silencing agents or immune-based therapies to determine whether it enhances the durability of viral suppression during ART interruption.

However, it is important to emphasize that the current analysis does not directly assess viral outcomes. Rather, it identifies systemic correlates that are hypothesized to support viral control based on prior mechanistic studies. Prospective trials in people living with HIV will be required to determine whether these proteomic changes translate into clinically meaningful effects on reservoir size, time to rebound, or functional cure.

### Alternative explanations and confounding

Several alternative explanations must be considered. First, confounding by indication remains a possibility, as metformin users differ systematically from non-users in ways that may influence inflammatory status. Although we attempted to mitigate this by comparing metformin users to diabetic individuals not receiving metformin and adjusting for key covariates such as age, sex, BMI, and socioeconomic status, residual confounding cannot be excluded.

Second, metabolic improvement itself may account for part of the observed effect. Metformin’s impact on glycemic control, insulin sensitivity, and adiposity could indirectly reduce inflammation, independent of direct immunologic mechanisms. The observation that BMI strongly influences certain inflammatory markers in our models supports the importance of metabolic context. Nevertheless, the persistence of significant associations after adjustment suggests that metformin exerts effects beyond simple metabolic normalization.

Third, reverse causation is unlikely but cannot be fully excluded in a cross-sectional design. Individuals with lower baseline inflammation may be more likely to tolerate or adhere to metformin therapy, although this is less plausible given prescribing patterns driven primarily by glycemic control.

In a large real-world analysis of patients with HIV, metformin use was associated with a significant reduction in all-cause mortality after rigorous propensity score matching (Figure 2, Table 5). The observed twofold higher mortality risk among patients not receiving metformin, together with the clear separation of Kaplan–Meier survival curves, suggests a clinically meaningful association that is unlikely to be explained solely by baseline differences in demographic or comorbidity profiles. The magnitude of the survival difference and early separation of Kaplan–Meier curves make it unlikely that residual confounding alone fully explains the observed association. These findings complement our population-scale proteomic analysis in the UK Biobank, in which metformin use was associated with reduced circulating levels of key inflammatory mediators, including interleukin-6 and CXCL10, as well as modulation of cytotoxic signaling pathways. Taken together, these results support a unified immunometabolic model in which metformin attenuates chronic systemic inflammation, thereby promoting a “cell-extrinsic” environment favorable for immune regulation and potentially improved viral control. Although causality cannot be established in this observational design, the concordance between mechanistic and clinical data strengthens the biological plausibility of a protective effect. Residual confounding, particularly related to diabetes severity and treatment selection, cannot be excluded, and the absence of HIV-specific clinical parameters such as viral load and CD4 count represents an additional limitation. Nonetheless, the magnitude and consistency of the association observed here support further investigation of metformin as a potential adjunctive therapy in HIV, particularly in the context of strategies aimed at reducing chronic immune activation and improving long-term outcomes.

### Limitations

This study has several important limitations. The observational, cross-sectional design precludes causal inference and limits our ability to establish temporal relationships between metformin exposure and proteomic changes. Longitudinal analyses would be valuable to determine whether initiation of metformin leads to dynamic shifts in inflammatory and cytotoxic markers over time.

In addition, the use of plasma proteomics provides only an indirect measure of immune activity. The “block-and-lock” mechanism operates primarily within tissue-resident CD4⁺ T cells and lymphoid compartments, which are not directly captured by circulating protein levels. While systemic markers such as IL-6 and CXCL10 are informative, they may not fully reflect the local microenvironment relevant to viral latency.

The relatively small number of individuals with HIV in the UK Biobank limits the ability to perform HIV-specific analyses. As a result, our conclusions are based on extrapolation from general population data combined with prior mechanistic insights. Dedicated cohorts of people living with HIV will be necessary to validate these findings in the target population.

Finally, while the Olink platform provides high-quality, reproducible measurements, it captures a predefined panel of proteins and may not encompass all relevant pathways. Additional multiomic layers, including transcriptomics and metabolomics, could provide a more comprehensive view of metformin’s effects.

### Future directions

Future studies should focus on integrating longitudinal and interventional designs to establish causality and clinical relevance. In particular, trials incorporating metformin into ART interruption protocols could directly test its impact on time to viral rebound.

Parallel mechanistic studies using primary human cells or tissue models could further delineate how metformin influences the balance between immune activation and viral latency.

Integration with genomic data may also identify subgroups of individuals who derive greater immunologic benefit from metformin, enabling precision approaches to adjunctive therapy. Finally, expanding analyses to include additional proteomic and metabolomic markers may uncover broader networks of immunometabolic regulation.

Prospective studies and randomized trials will be required to determine whether metformin directly improves survival or serves as a surrogate marker of broader metabolic and immunologic health in this population.

## Conclusion

In summary, this study provides large-scale evidence that metformin is associated with a coordinated reduction in systemic inflammation and modulation of cytotoxic immune activity. These findings support a model in which metformin promotes a “low-inflammatory” cell-extrinsic environment that may facilitate durable immune control of HIV. By bridging mechanistic insights from ATI cohorts with real-world population data, this work strengthens the rationale for evaluating metformin as a component of HIV cure strategies and highlights the broader potential of immunometabolic interventions in chronic viral disease.

## Data Availability

The data used in this study are available from the UK Biobank under approved application. UK Biobank data are not publicly available but can be accessed by qualified researchers upon application (https://www.ukbiobank.ac.uk
). The proteomic data utilized in this study were generated as part of the UK Biobank Pharma Proteomics Project using the Olink Explore platform.

